# Willingness to take the Covid-19 vaccines and associated factors at a tertiary institution community in Johannesburg, South Africa

**DOI:** 10.1101/2021.10.11.21264845

**Authors:** Bhadrashil H Modi, Deidré Pretorius, Joel M Francis

## Abstract

**Background:** SA is aiming to achieve herd immunity against Covid-19 by the first quarter of 2022. The success of the Covid-19 vaccination roll-out depends primarily on the willingness of the population to take the vaccines.

**Aim:** This study examined the willingness to take the Covid-19 vaccine, along with the factors of concern, efficacy, and preferences of the individual which may increase the willingness to be vaccinated.

**Setting:** This study was conducted at the University of the Witwatersrand, Johannesburg, amongst adult students and academic and professional staff.

**Methods:** We conducted a cross-sectional online study from 27 July – 14 August 2021. We performed descriptive and inferential analysis to determine the factors associated with willingness to take the Covid-19 vaccine.

**Results:** 2364 participants responded to a survey link and 82% were students, 66.8% were in the 18-29 years age band, females represented 64.0% and 49.2% were black people. 1965 (83.3%) were willing to receive a Covid-19 vaccine, the most preferred vaccines were Pfizer (41%) and J&J (23%), local pharmacy (29%) and GP (17%) were the preferred places for vaccination and the trusted sources of information on Covid-19 vaccines were the general practitioners (40.6%) and specialists (19,2%). Perceptions that vaccines are safe (aOR=31.56, 95%CI: 16.02-62.12 for affirmative agreement) and effective (aOR=5.92, 95%CI: 2.87-12.19 for affirmative agreement) were the main determinants for willingness to taking a COVID-19 vaccine

**Conclusion:** It is imperative to reinforce the message of Covid-19 vaccine safety and efficacy and to include the GPs and the community pharmacies in the vaccination roll-out in SA.

## Background

Covid-19 vaccination is the primary intervention, together with the other public health measures, in curbing the Covid-19 pandemic(1). The key success factor of the Covid-19 vaccination program is the willingness to be vaccinated, in addition to other factors such as effective communication and accessibility to the vaccines(2). The willingness to be vaccinated also influences the speed of the vaccination program, which is crucial as each vaccine administered implies fewer Covid-19 cases, hospitalisation, and death(3).

By mid-August 2021, 190 countries have begun mass vaccination campaigns and 4.7bn doses of the Covid-19 vaccines have been administered worldwide(4). In Europe and North America, where a large number of people have been vaccinated, there is a positive effect on reduced mortality, improved consumer, and business confidence, and thus the overall economy due to the opening of the countries(5). South Africa (SA) is aiming to vaccinate approximately 40m adults over the age of 18 years (67% of the population) to achieve herd immunity(6). To achieve this herd immunity by the first quarter of 2022, SA aims to vaccinate more than 200 000 people daily(7).

A global IPSOS survey undertaken in January 2021 in 15 countries showed that the population in the United Kingdom had the highest intent on being vaccinated (89%) and the lowest was in Russia (42%). The South African results showed a 61% intent on being vaccinated(8).

Evidence suggests that conspiracy beliefs and misinformation increase vaccine hesitancy and decrease the willingness to be vaccinated(9). It’s been reported that people who believe in such misinformation are four times less likely to be willing to be vaccinated and more likely to trust the information on social media.

The Wellcome global monitor 2018(10) reports that most people think vaccines are safe and effective. They further reported that almost 75% of the people, when it comes to health advice, would trust a doctor or nurse over anyone else (family, friends, leaders, or famous people) in deciding to be vaccinated or not. The Kaiser Family Fund (KFF) Covid-19 vaccine monitor (11) survey found that most people in the United States, turned to doctors, nurses, and other healthcare providers when deciding to be vaccinated. A follow-up survey found that 75% of people wish to obtain their Covid-19 vaccine at their doctor’s office, with 15% of the people chose pharmacies, followed by a hospital (9%)(12). Trust relationships exist not only in interpersonal relationships, such as the doctor-patient relationship but also with institutions. A trust-based relationship occurs when there is vulnerability on part of the truster, due to a knowledge imbalance and a lack of power within health institutions and the importance of one’s health and well-being(13).

There is limited evidence concerning the people’s willingness to take the Covid-19 vaccine in resource-constrained countries. In this study, we examined the willingness to take the Covid-19 vaccine, along with the factors of concern, efficacy, and preferences of the individual which may increase the willingness to be vaccinated.

## Methods

### Study design

The study was a cross-sectional online survey of adult students and staff members at the University of the Witwatersrand, Johannesburg, South Africa.

### Study population and setting

The study population was adults studying or working at the University of the Witwatersrand. All students and academic and professional staff older than 18 years were eligible to participate in the study. This university has a campus in Braamfontein and Parktown with approximately 40 129 registered students, 1 642 academic staff members, and nearly 3 960 administrative and service staff members.

### Sample size and sampling

Using the OpenEpi software (14) the sample was calculated based on the following assumptions, Wits community population (N=42,000), 50% of the participants will be willing to undertake COVID-19 vaccine, marginal error 5%, design effect of 2, and the 70% online survey nonresponse rate the estimated minimum sample size is 1300. We expected the distribution of participants to be at the ratio of 9:1 (1170 students and 130 staff). All adult students and staff were invited to participate in the study.

### Data collection procedures and ethical considerations

After obtaining ethics approval from the University of Witwatersrand Human Research Ethics Committee (M210495), and permission from the University of the Witwatersrand registrar, the registrar’s office sent the survey link to eligible students and staff. The email contained information on the study and participants had to consent before they could access the survey questionnaire. The questionnaire was based on the number of factors that will influence the successful and rapid roll-out of the Covid-19 vaccines, which includes the acceptability of the Covid-19 vaccine(15), the influence of social media(16), personal preferences(12), and accessibility and convenient sites that feels safe(17). The online survey ran from 27 July – 14 August 2021 and was closed thereafter. The survey responses were captured in Research Electronic Data Capture (REDCap) database (18). Data was stored in a password-protected cloud server. No identifying information was collected from the participants.

### Data management and analysis

Data from REDCap was exported to Stata version 14 for management and analysis. The dataset contained categorical variables and therefore we summarised the descriptive statistics frequencies (proportion). We used the chi-square test to determine the bivariate association of factors associated with willingness to undertake the COVID-19 vaccine. We decided a priori that age and gender will be forced in the multivariable logistic regression model regardless of the bivariate analysis p-value and other factors were entered into the logistic regression multivariable model if the bivariate analysis p-value was <0.20. We reported adjusted odds ratios and considered a p-value of <0.05 statistically significant.

## Results

### General characteristics of the study population

The university registrar shared the survey link with 40,129 students, 1,642 academic staff, and 3,960 professional and administrative staff. The survey response rate was 2364/45731 (5.0%). Table 1 shows that almost all participants responded to all survey questions 2330(98.6%). Among all participants (n=2364), 82% were students, 66.8% were in the 18-29 years age band, females represented 64.0% and 49.2% were black people.

**Table 1:** General characteristics of the study participants (N=2364)

| Characteristic | Categories | n | % |
| --- | --- | --- | --- |
| Completeness of the responses | Incomplete | 34 | 1.4 |
|  | Complete | 2330 | 98.6 |
|  | Total | 2364 | 100.0 |
| Age (Years) | 18-29 | 1580 | 66.8 |
|  | 30-49 | 571 | 24.2 |
|  | 50 and above | 213 | 9.0 |
|  | Total | 2364 | 100.0 |
| Gender | Male | 829 | 35.1 |
|  | Female | 1514 | 64.0 |
|  | Other | 21 | 0.9 |
|  | Total | 2364 | 100.0 |
| Highest level of education | Up to grade 12 | 732 | 31.0 |
|  | Diploma and undergraduate degree | 761 | 32.2 |
|  | Postgraduate degree | 867 | 36.7 |
|  | Total | 2360 | 100.0 |
| Race | Black | 1163 | 49.2 |
|  | White | 715 | 30.3 |
|  | Asian / Indian | 361 | 15.3 |
|  | Coloured | 103 | 4.4 |
|  | Other | 21 | 0.9 |
|  | Total | 2363 | 100.0 |
| Status (Position) at the university | Student | 1936 | 82.0 |
|  | Professional Staff | 138 | 5.8 |
|  | Academic Staff | 236 | 10.0 |
|  | Services | 50 | 2.1 |
|  | Total | 2360 | 100 |

### Willingness to receive COVID 19 vaccines and the preferred vaccines type

Table 2 further shows that 395/2360 participants (16.7%) indicated that they do not wish to receive COVID 19 vaccines. Among the 83.3% who indicated their willingness to take the COVID 19 vaccines - 30.7% had vaccinated (fully or partially). The most preferred COVID vaccine types by the participants were Pfizer (41.4%) and J&J (23.3%) vaccines. The least preferred ones were Sinovac (0.8%), AstraZeneca (1.9%), and Sputnik (1.8%). Among those willing to take the vaccines their main motivations were concern about the personal wellbeing (60.4%) and concerns of the wellbeing of others (61.3%). Side effects (58.0%) and short timelines of the vaccine trials (34.4%) were major concerns on the COVID 19 vaccines reported by the participants.

**Table 2:** Perceptions and willingness to take the COVID vaccines.

| Characteristic | Categories | n | % |
| --- | --- | --- | --- |
| COVID-19 vaccines are safe | Agree | 1134 | 48.1 |
|  | Somewhat agree | 842 | 35.7 |
|  | Somewhat disagree | 226 | 9.6 |
|  | Disagree | 157 | 6.7 |
|  | Total | 2359 | 100.0 |
| COVID-19 vaccines are effective | Agree | 1459 | 61.9 |
|  | Somewhat agree | 670 | 28.4 |
|  | Somewhat disagree | 121 | 5.1 |
|  | Disagree | 107 | 4.5 |
|  | Total | 2357 | 100.0 |
| What is your COVID-19 vaccination status | Not eligible as yet | 969 | 41.1 |
|  | Registered, but not vaccinated | 272 | 11.5 |
|  | Vaccinated - partially | 302 | 12.8 |
|  | Vaccinated - fully | 422 | 17.9 |
|  | Do not wish to be vaccinated | 395 | 16.7 |
|  | Total | 2360 | 100.0 |
| What is your preferred COVID-19 vaccine | J & J | 549 | 23.3 |
|  | Pfizer | 977 | 41.4 |
|  | Sputnik | 42 | 1.8 |
|  | Astra Zeneca | 45 | 1.9 |
|  | Sinovac | 18 | 0.8 |
|  | Does not matter which one | 430 | 18.2 |
|  | None | 298 | 12.6 |
|  | Total | 2359 | 100.0 |
| Concerns about COVID-19 vaccines (n=2364)<sup>1</sup> | No Concern | 722 | 30.5 |
|  | Worried about side effects | 1372 | 58.0 |
|  | Vaccine trials were too fast | 813 | 34.4 |
|  | Risk of getting COVID is low | 140 | 5.9 |
|  | Vaccines are not effective | 311 | 13.2 |
|  | I am against vaccines in general | 92 | 3.9 |
|  | Other reasons | 190 | 8.0 |
|  | Total | 2364 |  |
| Reasons for taking the COVID-19 vaccine(n=2364)<sup>1</sup> | Will not take the vaccine | 324 | 13.7 |
|  | Concerned of personal wellbeing | 1428 | 60.4 |
|  | Concerned of wellbeing of others | 1450 | 61.3 |
|  | It is the right thing to do | 1141 | 48.3 |
|  | Fear of dying | 504 | 21.3 |
|  | Travelling | 767 | 32.4 |
|  | Pressure from employer | 152 | 6.4 |
|  | Other reasons | 110 | 4.7 |

<sup>1</sup>Each category was a standalone question

### Perceptions on safety and effectiveness of the COVID-19 vaccines

Of the participants (n=2359) who responded to the question on the safety of COVID 19 vaccines, 1134(48.1%) agreed affirmatively that the vaccines were safe and 35.7% of the respondents somewhat agreed that the vaccines were safe. On the other hand, 2357 participants who responded to the vaccine effectiveness question,1459(61,9%) agreed affirmatively that the COVID vaccines were effective and 28.4% indicated that they somewhat agreed that the vaccines are effective (Table 2).

### Preferred COVID 19 vaccinations places and the sources of COVID 19 information

Table 3 of the results describe the preferred places for vaccination and sources of information. Participant’s preferences were a local pharmacy (28.8%), general practitioner (17.2%), hospital (9.9%), local health clinic (8.3%), workplace (7.7%), and vaccination site run by the government (7.8%). The trusted sources of information on Covid-19 vaccines were the general practitioners (40.6%), specialists (19,2%), the Government/Department of Health (12.6%), and the Medical Aids (6.7%). The main sources of information reported were by the Government/Department of Health (60.3%), via mainstream media (Radio, TV, Newspapers) (56.4%), social media (48.4%) and by Medical Aids (26.0%).

**Table 3:**
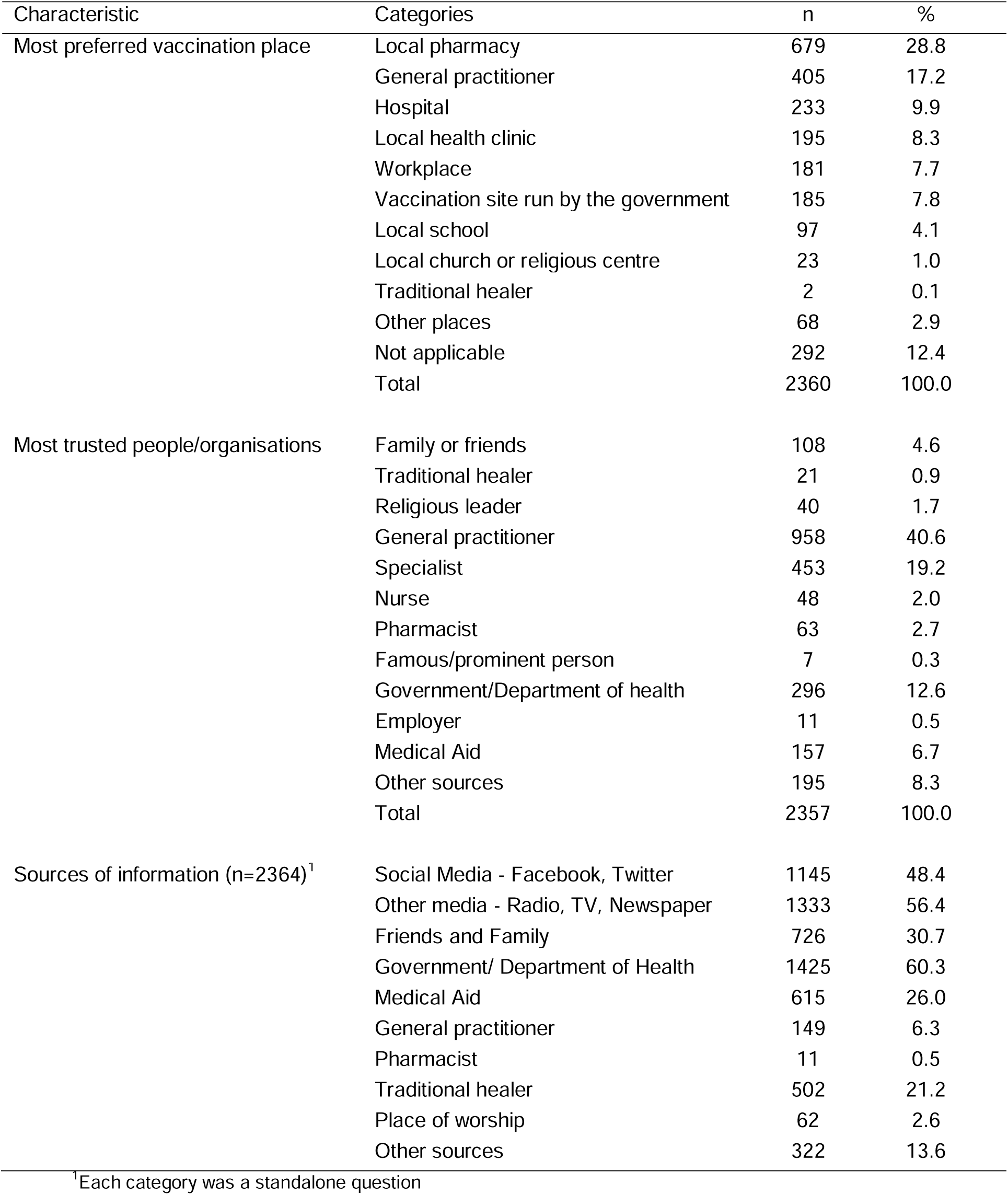
Preferred vaccination place and sources of information for COVID 19 vaccines.

| Characteristic | Categories | n | % |
| --- | --- | --- | --- |
| Most preferred vaccination place | Local pharmacy | 679 | 28.8 |
|  | General practitioner | 405 | 17.2 |
|  | Hospital | 233 | 9.9 |
|  | Local health clinic | 195 | 8.3 |
|  | Workplace | 181 | 7.7 |
|  | Vaccination site run by the government | 185 | 7.8 |
|  | Local school | 97 | 4.1 |
|  | Local church or religious centre | 23 | 1.0 |
|  | Traditional healer | 2 | 0.1 |
|  | Other places | 68 | 2.9 |
|  | Not applicable | 292 | 12.4 |
|  | Total | 2360 | 100.0 |
| Most trusted people/organisations | Family or friends | 108 | 4.6 |
|  | Traditional healer | 21 | 0.9 |
|  | Religious leader | 40 | 1.7 |
|  | General practitioner | 958 | 40.6 |
|  | Specialist | 453 | 19.2 |
|  | Nurse | 48 | 2.0 |
|  | Pharmacist | 63 | 2.7 |
|  | Famous/prominent person | 7 | 0.3 |
|  | Government/Department of health | 296 | 12.6 |
|  | Employer | 11 | 0.5 |
|  | Medical Aid | 157 | 6.7 |
|  | Other sources | 195 | 8.3 |
|  | Total | 2357 | 100.0 |
| Sources of information (n=2364)<sup>1</sup> | Social Media - Facebook, Twitter | 1145 | 48.4 |
|  | Other media - Radio, TV, Newspaper | 1333 | 56.4 |
|  | Friends and Family | 726 | 30.7 |
|  | Government/ Department of Health | 1425 | 60.3 |
|  | Medical Aid | 615 | 26.0 |
|  | General practitioner | 149 | 6.3 |
|  | Pharmacist | 11 | 0.5 |
|  | Traditional healer | 502 | 21.2 |
|  | Place of worship | 62 | 2.6 |
|  | Other sources | 322 | 13.6 |

<sup>1</sup>Each category was a standalone question

### Factors associated with willingness to take the COVID 19 vaccines

Overall, the prevalence of willingness to take the COVID vaccines was 83.3%. All reported sociodemographic factors, the perceptions of vaccine safety and effectiveness, the perception that one was at low risk of COVID infection, and generally being against all vaccines were associated with the willingness to take the COVID 19 vaccines at the bivariate analysis (Table 4). However, only the perception that the COVID 19 vaccines are safe (aOR=31.56, 95%CI: 16.02-62.12 for affirmative agreement and aOR=9.38, 95%CI: 5.40-16.30 for a somewhat agreement) and that the vaccines are effective (aOR=5.92, 95%CI: 2.87-12.19 for affirmative agreement, and aOR=2.90, 95%CI: 1.48-5.65 for a somewhat agreement) remained significantly associated with the willingness to take COVID 19 vaccines.

**Table 4:** Bivariate analysis of factors associated with willingness to take COVID-19 vaccines.

| Characteristic | Categories | Willing to take COVID 19 vaccines |  |  |  | P value<sup>1</sup> |
| --- | --- | --- | --- | --- | --- | --- |
|  |  | No |  | Yes |  |  |
|  |  | n | % | n | % |  |
| Age | 18-29 | 290 | 18.4 | 1287 | 81.6 | <0.001 |
|  | 30-49 | 92 | 16.1 | 478 | 83.9 |  |
|  | 50 and above | 13 | 6.1 | 200 | 93.9 |  |
|  | Total | 395 | 16.7 | 1965 | 83.3 |  |
| Gender | Male | 151 | 18.2 | 677 | 81.8 | 0.138 |
|  | Female | 243 | 16.1 | 1268 | 83.9 |  |
|  | Other | 1 | 4.8 | 20 | 95.2 |  |
|  | Total | 395 | 16.7 | 1965 | 83.3 |  |
| Highest level of education | Up to grade 12 | 146 | 20.0 | 584 | 80.0 | <0.001 |
|  | Diploma and undergraduate degree | 135 | 17.8 | 625 | 82.2 |  |
|  | Postgraduate degree | 111 | 12.8 | 755 | 87.2 |  |
|  | Total | 392 | 16.6 | 1964 | 83.4 |  |
| Race | Black | 304 | 26.2 | 856 | 73.8 | <0.001 |
|  | White | 43 | 6.0 | 672 | 94.0 |  |
|  | Asian / Indian | 32 | 8.9 | 328 | 91.1 |  |
|  | Coloured | 14 | 13.6 | 89 | 86.4 |  |
|  | Other | 1 | 4.8 | 20 | 95.2 |  |
|  | Total | 394 | 16.7 | 1965 | 83.3 |  |
| University status(position) | Student | 360 | 18.6 | 1573 | 81.4 | <0.001 |
|  | Professional Staff | 15 | 10.9 | 123 | 89.1 |  |
|  | Academic Staff | 10 | 4.2 | 226 | 95.8 |  |
|  | Services | 9 | 18.4 | 40 | 81.6 |  |
|  | Total | 394 | 16.7 | 1962 | 83.3 |  |
| Perceiving COVID 19 vaccines are safe | Agree | 31 | 2.7 | 1103 | 97.3 | <0.001 |
|  | Somewhat agree | 113 | 13.4 | 728 | 86.6 |  |
|  | Somewhat disagree | 127 | 56.2 | 99 | 43.8 |  |
|  | Disagree | 123 | 78.3 | 34 | 21.7 |  |
|  | Total | 394 | 16.7 | 1964 | 83.3 |  |
| Perceiving COVID 19 vaccine are effective | Agree | 69 | 4.7 | 1390 | 95.3 | <0.001 |
|  | Somewhat agree | 168 | 25.1 | 501 | 74.9 |  |
|  | Somewhat disagree | 70 | 57.9 | 51 | 42.1 |  |
|  | Disagree | 86 | 80.4 | 21 | 19.6 |  |
|  | Total | 393 | 16.7 | 1963 | 83.3 |  |
| Perceiving risk of getting COVID is low | No | 362 | 16.3 | 1858 | 83.7 | 0.026 |
|  | Yes | 33 | 23.6 | 107 | 76.4 |  |
|  | Total | 395 | 16.7 | 1965 | 83.3 |  |
| Being against vaccines in general | No | 336 | 14.8 | 1932 | 85.2 | <0.001 |
|  | Yes | 59 | 64.1 | 33 | 35.9 |  |
|  | Total | 395 | 16.7 | 1965 | 83.3 |  |

<sup>1</sup> p-value based on the Chi-Square test

**Table 5:** Factors associated with willingness to take COVID 19 vaccines.

| Characteristic | Categories | Willing to take COVID 19 vaccine |  |  |  |
| --- | --- | --- | --- | --- | --- |
|  |  | Yes | %<sup>1</sup> | Adjusted OR | 95% CI |
| Age | 18-29 | 1287 | 81.6 | 1 |  |
|  | 30-49 | 478 | 83.9 | 1.20 | 0.78 - 1.83 |
|  | 50 and above | 200 | 93.9 | 1.99 | 0.88 - 4.51 |
| Sex<sup>2</sup> | Male | 677 | 81.8 | 1 |  |
|  | Female | 1268 | 83.9 | 1.28 | 0.96 - 1.73 |
| Highest level of Education | Up to grade 12 | 584 | 80.0 | 1 |  |
|  | Diploma and undergraduate degree | 625 | 82.2 | 1.27 | 0.91 - 1.79 |
|  | Postgraduate degree | 755 | 87.2 | 1.06 | 0.71 - 1.60 |
| Race | African | 856 | 73.8 | 1 |  |
|  | White | 672 | 94.0 | 1.31 | 0.86 - 1.98 |
|  | Asian / Indian | 328 | 91.1 | 1.61* | 1.01 - 2.56 |
|  | Coloured | 89 | 86.4 | 1.43 | 0.69 - 2.98 |
|  | Other | 20 | 95.2 | 1.31 | 0.15 - 11.62 |
| University status(position) | Student | 1573 | 81.4 | 1.53 | 0.56 - 4.18 |
|  | Professional Staff | 123 | 89.1 | 3.60* | 1.10 - 11.75 |
|  | Academic Staff | 226 | 95.8 | 2.82 | 0.83 - 9.63 |
|  | Services | 40 | 81.6 | 1 |  |
| Perceiving COVID 19 vaccines are safe*** | Agree | 1103 | 97.3 | 31.56*** | 16.04 - 62.12 |
|  | Somewhat agree | 728 | 86.6 | 9.38*** | 5.40 - 16.30 |
|  | Somewhat disagree | 99 | 43.8 | 1.87* | 1.07 - 3.26 |
|  | Disagree | 34 | 21.7 | 1 |  |
| Perceiving COVID 19 vaccine are effective*** | Agree | 1390 | 95.3 | 5.92*** | 2.87 - 12.19 |
|  | Somewhat agree | 501 | 74.9 | 2.90** | 1.48 - 5.65 |
|  | Somewhat disagree | 51 | 42.1 | 1.83 | 0.88 - 3.80 |
|  | Disagree | 21 | 19.6 | 1 |  |
| Perceiving risk of getting COVID is low | No | 1858 | 83.7 | 1 |  |
|  | Yes | 107 | 76.4 | 0.85 | 0.50 - 1.43 |
| Being against vaccines in general | No | 1932 | 85.2 | 1 |  |
|  | Yes | 33 | 35.9 | 0.62 | 0.35 - 1.09 |

\*\*\* p<0.001, \*\* p<0.01, \* p<0.05
<sup>1</sup>Row percentage of Yes responses across each category,
<sup>2</sup>excluded the other gender because of the small sample

## Discussion

Key findings of this study were the more than 80% of participants declared their willingness to be vaccinated. They also had a clear preference for Pfizer and Johnson and Johnson vaccines. Mainstream media and social media were sources of information and safety and efficacy were prominent determinants of willingness to vaccinate. The trusted sources of information on Covid-19 vaccines were the general practitioners (40.6%), specialists (19,2%).

The findings of this study are consistent with the result of the Coronavirus Rapid Mobile Survey(19), which found, in May 2021, that 76% of adults in South Africa are willing to take the Covid-19 vaccine. In a global survey of acceptance of the Covid-19 vaccine, China had the highest positive responses (86%), Russia had the lowest positive responses (54.9%). South Africa (81.58%) fell in the group of the high tendency towards acceptance in middle-income countries, together with Brazil and India(20).

Several studies have found a 3-way relationship between the factors of perceived severity, perceived susceptibility, and perceived benefits which influenced the willingness to take the Covid-19 vaccine (2,21,22). Although most of the study population felt these vaccines were safe and effective, the concerns around side effects and the rapid clinical trials to test these vaccines are possible barriers; the primary motivation to take the vaccine included personal well-being, as well as the well-being of others. Thus, these factors meet two criteria of the triad (perceived susceptibility and perceived benefits) and will influence their willingness to take the vaccine.

The willingness may also have been influenced in that the survey included participants in a tertiary education facility with better access to information and developed reasoning skills. A study in Thailand showed that increased health education increased willingness in the uptake of influenza vaccinations(23), while a study in America suggested that people who were more knowledgeable about herd immunity, were also more willing to take vaccinations(24). A study in Nigeria found a high level of hesitancy amongst medical and nursing students, however marginally more under nurses, who had lower risk perception and knowledge of Covid-19(25).

Vaccine hesitancy is an important hurdle in the willingness to be vaccinated, thus, hesitancy is an essential component to address for a successful vaccination rollout(26). Razia and colleagues(27) proposed a 5-C model to influence vaccine hesitancy. This model consists of ‘Confidence (importance, safety and efficacy of vaccines); Complacency (perception of low risk and low disease severity); Convenience (access issues dependent on the context, time and specific vaccine being offered); Communications (sources of information); and Context (sociodemographic characteristics)’.

In this study, although the main sources of health information concerning the Covid-19 vaccines were obtained via the department of health, social media (Facebook, Twitter, Instagram), mainstream media (radio, television, and newspapers), and the medical aids, the most trusted source of the health information was from general practitioners and followed by specialists. This is new information in the South African context. Although many study participants obtained their vaccine-related information from social media, the study participants based in a tertiary institution seem discerning in interpreting the information and not be swayed by the conspiracy theories that abound on social media(9).

The trusted source of information result is also in keeping with Opel et al (28), who found that a child’s healthcare provider is a trusted resource in the parental decision to vaccinate their child. In vaccine-hesitant parents living in the United States of America, they cite the child’s healthcare provider as the key to changing their minds.

Most of the study population were keen on obtaining two types of Covid-19 vaccines currently offered in SA. Willingness to vaccinate may decrease if the other vaccine products become available in the market and people do not have a choice of the vaccine they will receive.

In countries where the Covid-19 vaccination has been successfully rolled out to the majority of the population, primary healthcare and community-based teams played a central role in rolling out the vaccination program(29–31). In this study, to increase the willingness to be vaccinated, most people preferred being vaccinated at their local pharmacy and their general practitioner, as opposed to large vaccination sites which currently exist.

The findings of this study should be interpreted considering the following limitations. First, a poor response rate (5%) introduces the selection bias of participation of those using the university email, internet access (for those working from home), and those interested in the topic. Second, a potential risk to a social desirability response of over-reporting willingness to take the COVID-19 vaccines but the online anonymous survey would have mitigated this bias. Third, even though the university community is comprised of individuals from all walks of life and geographical areas in the Gauteng province, the findings may not be entirely generalizable to the general population but university communities in South Africa. Finally, because of the cross-sectional nature of the study, we are only able to determine the factors associated with willingness to take the COVID-19 vaccine.

## Conclusion

The current study identified that the safety and efficacy of the Covid-19 vaccines are significantly associated with the willingness to be vaccinated against COVID-19. There is an urgent need to reinforce the communication around vaccines safety and effectiveness for the individual and the community, through doctors, media, and other approaches to reduce vaccine hesitancy and increase the willingness to be vaccinated. The policymakers should consider incorporating the community pharmacies and the GPs in the primary care-led vaccination roll-out, in addition to the current large vaccination sites, to achieve herd immunity as soon as possible.

## Data Availability

All data produced in the present study are available upon reasonable request to the authors

## Author contribution

All the authors contributed to the design and implementation of the research, to the analysis of the results and the writing of the manuscript.

## Data availability

The data that support the findings of this study are available from the corresponding author, [BM], upon reasonable request.

## Disclaimer

The views expressed in the submitted article are the author’s own and not an official position of the institution.

